# Reduced Mortality During Holidays and the COVID-19 Pandemic in Israel

**DOI:** 10.1101/2020.07.16.20155259

**Authors:** Roni Rasnic, Danielle Klinger, Dan Ofer, Yoav Comay, Michal Linial, Eitan Bachmat

**Affiliations:** The Rachel and Selim Benin School of Computer Science and Engineering, The Hebrew University of Jerusalem, Israel; Department of Biological Chemistry, Institute of Life Sciences, The Hebrew University of Jerusalem, Jerusalem, Israel; Faculty of Health Science, School of Medicine, Ben Gurion University, Beer-Sheva, Israel; Department of Computer Science, Ben-Gurion University, Beer-Sheva, Israel

**Keywords:** Epidemiological reports, Nosocomial infections, natural death, multivariable analysis

## Abstract

Evidence suggests varied trends in mortality surrounding the holiday period. Most studies support an association between increased mortality rates and holidays. We compare the effect of the number of holiday days per week on the overall mortality rate in the Israeli population. Between 2000-2020, we see significantly reduced mortality rates in weeks containing national holidays. We observed the same trend in all-cause mortality during the 3-weeks COVID-19 pandemic lockdown. As the Israeli health care system, and hospitals especially, function near peak capacity year-round, we propose that reduced medical service utilization during holidays and the COVID-19 lockdown period might underlie the lower mortality rates.

## Introduction

The health care system in each country operates at a reduced capacity over weekends and holidays. Several studies were conducted in different countries and communities to assess the impact of weekends and holidays on all-cause mortality (1-5). We tested whether restricted hospital services during holidays in Israel have any impact on all-cause mortality.

Ample studies show variable results regarding the impact of major holidays, and the period around them on the national death toll (6-8). In southern England, no effect on mortality was apparent within 2-days from weekends. However, respiratory deaths accelerated sharply at Christmas week (9). In Denmark, despite a decrease in the admission rates in weekend days, an observed increase in mortality was reported (10). In the USA among patients hospitalized with acute kidney injury, weekend admission is associated with a higher risk for death (11). In Canada and France, stroke patients admitted on weekends had higher risk-adjusted mortality compared to patients admitted on weekdays (12, 13). A large-scale study found that cancer patients are especially vulnerable to the effects of holidays and weekends. Indeed, cancer patients show higher mortality relative to patients admitted on weekdays (14).

Notably, mortality during holidays and weekends were also affected by cultural and social norms (15). In South Korea, the incidence of out-of-hospital cardiac arrest occurred more frequently on traditional holidays than on other day types (e.g. weekend, weekday, public holidays) (16). In addition, the unique emotional impact resulted from traditional and major religious holidays may increase or attenuate mortality (4). The proposed beneficial effect of some traditional holidays of delayed death had not been validated (e.g., (17)).

Increased mortality rates during weekends and holidays are typically attributed to the reduction in medical services - both elective and emergency, as well as lack of compliance with medication or greater difficulty in getting medical help. The overindulgence or the stress of the holiday might also contribute to excess deaths during the holidays (10, 18-20). The impact on mortality was also quantified in unplanned events such as pandemic event. For example, the mortality caused by diabetes mellitus and cerebrovascular diseases significantly increased during the SARS epidemic (21)

In Israel, accessibility for hospital services is markedly reduced during national holidays. During holidays the outpatient clinics and hospitals are operating at a reduced capacity (i.e., emergency-mode). Furthermore, during holidays, public transportation and municipal services are minimal, thus affecting acute medical service accessibility. The Israeli Semitic holidays offer a unique perspective for testing the phenomenon of altered mortality during holidays. Specifically, Israeli holidays are based on a lunar cycle and occur during different weeks when aligned to the Gregorian civil calendar and start on varying weekdays. We compare the effect of the number of holiday days per week on the overall mortality rate in the Israeli population over the past 20 years. We also focus on changes in all-cause mortality during the 3-week COVID-19 lockdown in Israel. We examined whether significant differences in the number of deaths in Israel during holidays exist. We hypothesize that reduced medical service utilization during national holidays and the COVID-19 lockdown period explains the changes in Israel’s overall mortality rates.

## Methods

The data consists of a weekly count of all-cause mortality in the population of Israel between January 2000 and May 2020. This includes the 2020 COVID-19 pandemic months, and the mandatory national lockdown (weeks 12-14). The data was acquired from weekly epidemiological reports of the Israeli ministry of health (MOH). The data was aligned with the Gregorian calendar (1-53 weeks). Weekly deaths were normalized by the population size. Israeli holiday dates were inserted and the total number of holiday days were summed per week, varying between 0-2.5 (holiday half-days were counted as 0.5). Holidays that landed on Saturdays weren’t included in the summed total of holiday days per week. The mean weekly mortality rate in weeks with holidays, in comparison with holiday-free weeks, was examined in a two-sided t-test.

We measured the effect of the number of vacation days on weekly mortality in a univariate linear regression model. We examined the mean weekly mortality rate during COVID-19 lockdown weeks, in comparison to the aligned holiday-free weeks. Lastly, we conducted multivariable regression using the year and week number as covariates. Analyses were performed using R.

## Results

Israel has 8 national holidays a year. On average there are 12 national holiday days a year (not including weekend day. Between 2000 and 2020 we observe a significantly lower all-cause mortality rate in weeks containing national holidays in comparison with holiday-free weeks, with a p-value of 1.5e-13 (**Fig 1A**). Mean weekly mortality rate distribution analyzed according to the number of additional holiday days each week also showed a significant difference, (p-value of p=2.2e-16, **Fig 1B**). We further analyzed mortality in weeks covering the COVID-19 lockdown. The mean weekly mortality rate was significantly lower in weeks covering the COVID-19 lockdown concerning the same holiday-free weeks, p=2.9e-9 (**Fig 1C**).

**Fig 1.**
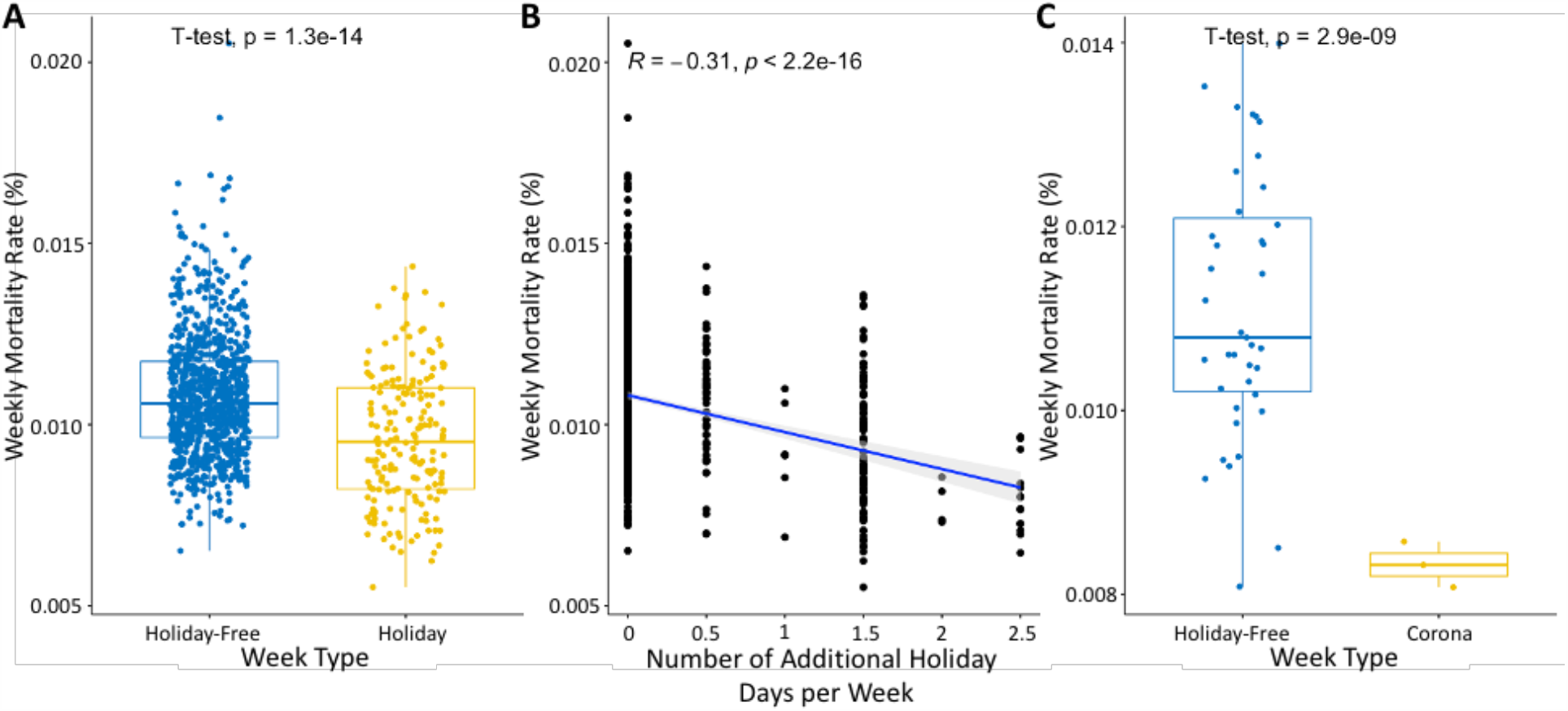
Statistical analysis of mortality rate in COVID-19 pandemic, holiday-free, and holiday weeks. **(A)** Mean weekly all-cause mortality rate in holiday-free (blue) and holiday (yellow) weeks. **(B)** Correlation of the number of additional holiday days per week with the weekly mortality rate. The shaded area represents the 95% confidence interval. **(C)** Mean weekly all-cause mortality rate in holiday-free (blue) and COVID-19 lockdown (yellow) weeks (weeks 12-14 scaled to the Gregorian calendar).

Applying linear regression showed that ‘holiday weeks’ is the coefficient with the strongest effect (beta coefficient) and second most significant p-value, second to the year coefficient (Estimate = −8.43; p = 3.74E-18, **Fig 2**).

**Fig 2.**
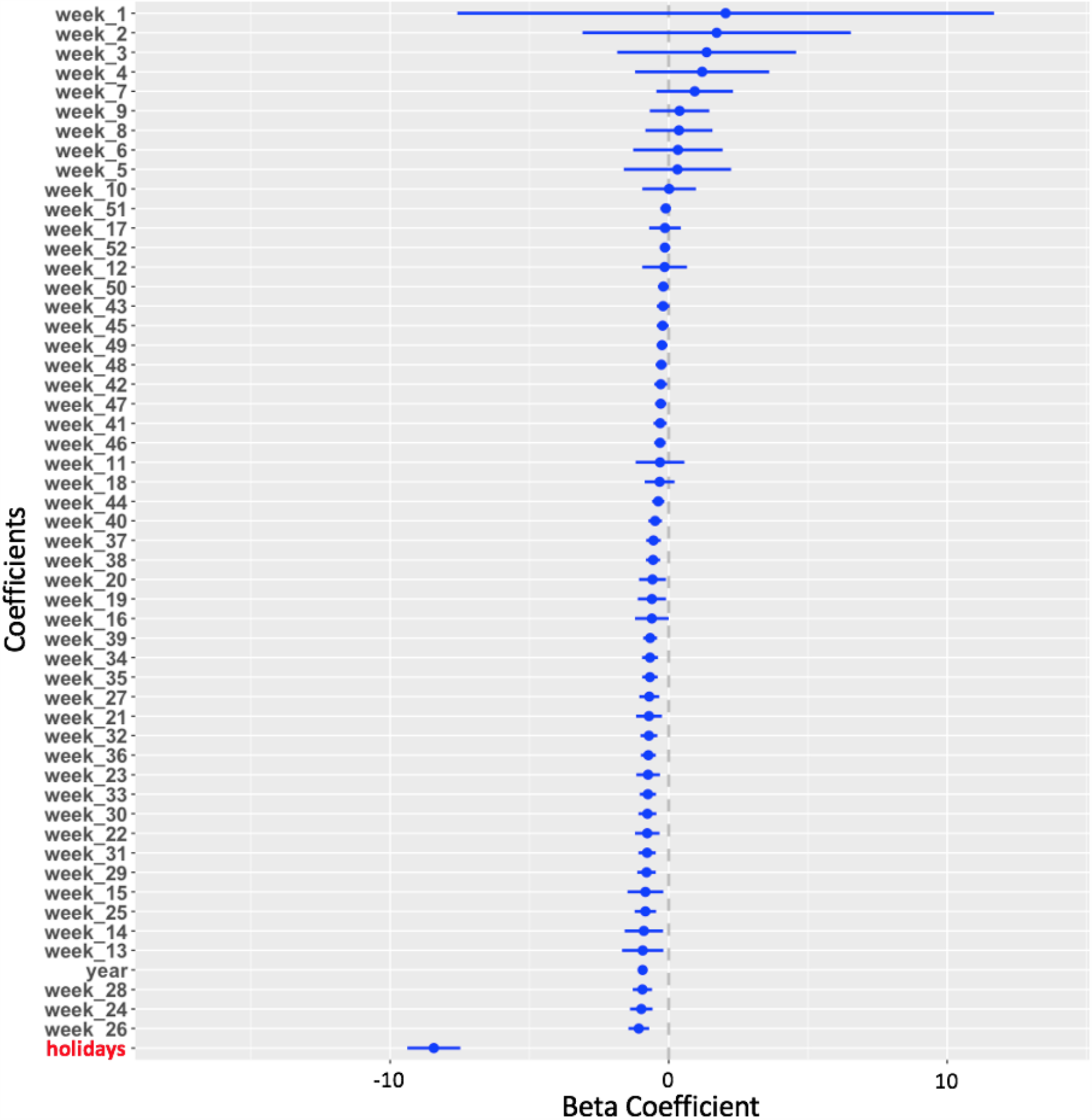
Multivariable linear regression analysis. Beta coefficients of the normalized lasso regression for the weekly mortality rate are shown. Blue lines represent the coefficients’ 95% confidence intervals. Note that the coefficients for ‘year’ and ‘holidays’ had the most significant estimates (p-value = 5.808e-39; p-value = 3.742e-18, respectively). Holiday days varied between 8.5-14 per year, spanning over 9-10 weeks a year.

## Discussion

Our findings suggest a significant difference in overall mortality in the Israeli population between weeks with and without holidays. Also, during the COVID-19 lockdown, hospitals in Israel switched to a ‘holiday routine’ mode and operated at a reduced capacity. We observed the same phenomenon of reduced mortality during these weeks as during holiday periods in the preceding 20 years (Fig. 1). Importantly, holidays in Israel are associated with cross-generation social gatherings (e.g., family, prayer), but such gatherings were strongly suppressed and unauthorized during the COVID-19 pandemic lockdown (22). The inverse trend in personal interactions allows us to examine competing hypotheses regarding the mortality rates.

Israel, similar to many other European countries, has a subsidized national healthcare system with full coverage. Therefore, the urgency for an acute hospitalization is not governed by immediate budgetary considerations. During the holidays and the COVID-19 lockdown, both elective and emergency medical procedures are reduced dramatically. An alternative to the accepted view that a reduction in medical services during weekends and holidays-increases death rates, due to postponing urgent services, we hypothesize that the imposed reduction in medical services during holidays and unplanned events (COVID-19 lockdown, labor strike of medical stuff) allowed improved management of overloaded hospitals and healthcare professionals, and a cumulative reduction in all-cause death in Israel.

The Israeli health care system is regularly at over-capacity year-round (94% occupancy, with 2.2 beds per 1000 people, versus the OECD average of 77% and 64% in the USA (23). The difference is extended also to practicing physicians with 3.3 relative to 4.8 per 1000 population in Israel and the OECD, respectively (23). The number of nurses in Israel is only 54% of that of the OECD. It was reported by the ministry of health (MOH) that in Israel, some 4,000-6,000 annual deaths are attributed to nosocomial infections (NI). Such health-care infections are a common cause of mortality in hospitals worldwide (24). According to the World Health Organization (WHO), about 15% of hospitalized patients will suffer such infections (25), and 1 out of 17 will die (24). Based on the Israeli ministry of health (MOH report, years 2011-2016) septicemia as a cause of death has been increasing in recent years in Israel. This is likely to reflect physical conditions (i.e. bed density, ventilation), the use of invasive devices, and more. Notably, In Israel death from sepsis was ranked as the 5th top cause of death, while it was not amongst the 10 leading causes of death in most countries. Altogether we present a likely shared explanation for the all-cause mortality trend of holidays and the COVID-19 lockdown which concerns the significant reduction in medical service utilization. Presumably, reduced hospital utilization allows improved management and reduction in workload and overall improved personal care.

## Conclusions

We present observational evidence covering >20 years in Israel. The consistent observation for the COVID-19 lockout and the year-around national holidays supports the need for diffused and sparse medical services and the expected benefit from strengthening community and home medical services.

## Data Availability

Date are public and can be share upon need

## Acknowledgements

We thank Nadav Rappoport and the Linial’s lab for useful discussion.

